# Brain health Begins Before Birth (B4): A “learning” pregnancy and birth cohort study

**DOI:** 10.1101/2025.03.12.25323835

**Authors:** Rachel L. Pride, Jannely Villarreal, Daniela Restrepo, Victoria A. Grunberg, Maria Bazan, Devika Subedi, Allison Fitzgerald, Katherine Shauh, Elena Wollman, Sofia Perdomo, Oren M. Bazer, Sharon Dekel, Cindy H. Liu, Rakesh Karmacharya, Julie H. Levison, Paul H. Lerou, Erin C. Dunn, Joshua L. Roffman

## Abstract

Mental health disorders affect over 1 billion people worldwide, profoundly impacting individuals, families, and the global economy. Risk for psychopathology begins early in life, with the perinatal period representing a critical window of vulnerability. The Brain health Begins Before Birth (B4) Study at Massachusetts General Hospital (MGH) is a prospective birth cohort designed to identify modifiable risk and resilience factors influencing early brain development and child- and adult-onset psychopathology. This study aims to deeply phenotype prenatal exposures through maternal surveys administered at multiple time points during and after pregnancy; assess early neurodevelopmental outcomes through the first two years of life; and triangulate exposure and outcome data with underlying biological mechanisms. The B4 Study consists of structured, remote surveys covering psychosocial, environmental, and health-related factors throughout pregnancy and the first two years of life, supplemented by medical record review. By integrating risk and resilience factors into a dynamic learning cohort model, the B4 Study aims to advance the field of preventive psychiatry by identifying actionable pathways for early intervention, testing strategies in real-time, and ultimately shaping policies that promote lifelong mental health and wellbeing from the earliest stages of development.

## Introduction

Mental health disorders account for at least 7% of the global burden of disease, affecting over 1 billion people worldwide.^1–4^ In the United States, this includes nearly one in five adults, one in six children aged 6-17, and almost half (49.5%) of adolescents aged 13-18.^5,6^ Without proper treatment, mental illness can inflict significant strains on the quality of life and socioeconomic well-being of entire populations, comprising around 15% of all global years lived with disability and costing the global economy an estimated $2.5 trillion per year.^2,4^ Risk for mental health disorders increases significantly during pregnancy and the first year postpartum (the perinatal period).^7,8^ In addition, mental illness in early life has been associated with decreased social and economic mobility in adulthood, as well as premature mortality.^9,10^ However, fewer than half of U.S. children and adults living with mental illness receive treatment, and access to care is disparate across socioeconomic, geographic, racial, and ethnic lines.^5,11–14^

It has been well-established that risk for mental illness originates years before onset and is influenced by various factors throughout the life course, ranging from insecure early childhood attachment to financial adversity to acute physical trauma.^15–18^ While many genetic, environmental, and demographic risk and resilience factors are relatively fixed, modifiable factors such as smoking, parenting style, and early life nutrition have also been shown to influence psychopathology risk.^19–21^ Thus, many efforts to advance mental health equity have shifted focus to prevention, aiming to alleviate the risk for mental illness prior to onset, as well as promote resilience factors believed to improve outcomes.^6,22–24^

The theory of developmental origins of health and disease, or the Barker hypothesis, emerged nearly 40 years ago, suggesting a link between prenatal nutrition and risk for coronary heart disease based on epidemiological studies in England and Wales.^25–27^ In the years since, prenatal programming of various cardiovascular health outcomes, such as metabolic syndrome, hypertension, type 2 diabetes, and stroke, has been studied extensively.^28–31^

Converging evidence implicates similar prenatal programming mechanisms in risk for psychiatric and neurodevelopmental disorders such as attention-deficit hyperactivity disorder, autism, and schizophrenia, among others.^3,17,22,28,32–36^ Previously identified prenatal risk factors for increased psychopathology risk include maternal infection, stress, substance use, and obstetric complications.^3,21,33,34,37,38^ Our recent analysis of data from the Adolescent Brain and Cognitive Development (ABCD) study suggests that elevated psychopathology risk due to adverse prenatal exposures may be dose-dependent, showing a linear increase of behavioral symptoms at age 9-10 with each added exposure.^33^

In the context of the COVID-19 pandemic, this work has taken on added urgency. For one, direct maternal infection with SARS-CoV-2 may increase the risk for maternal and neonatal morbidity and mortality, including preeclampsia/eclampsia, preterm birth, and ICU/NICU admission.^39,40^ After recovery, a constellation of symptoms and conditions referred to as post-acute sequelae of COVID-19 (PASC), or long COVID, can include increased symptoms of anxiety, depression, and PTSD, as well as cognitive deficits.^41–44^ In addition, the pandemic itself exacerbated many existing perinatal risk factors for child- and adult-onset psychopathology, including psychological distress, alcohol and substance use, and social isolation.^45–49^

Conversely, specific behaviors during pregnancy have been identified as potentially protective against adverse neurodevelopmental and psychiatric outcomes, namely exercise, physical activity, nutrition, and periconceptional folic acid supplementation.^3,50,51^ Early life resilience factors such as secure attachment, positive parental affect, and high-quality social support/environment have also been suggested to mitigate psychopathology risk.^21,52–54^

Identification and further clarification of risk factors remain top priorities in the field of preventive psychiatry, inspiring new birth cohort studies such as the HEALthy Brain and Child Development Study (HBCD) and new analyses of existing birth cohort data such as the 1987 Finnish Birth Cohort (FBC).^55,56^ However, characterizations of such risk are seldom coupled with analyses of resilience factors, leaving little to be done in the case of unmodifiable risk factors. While an increasing number of studies have highlighted the importance of prenatal and early life intervention for the prevention of mental illness, few if any studies have attempted to incorporate such interventions into the fabric of a prospective birth cohort study, and none have focused specifically on resilience factors.^57–59^ The Brain health Begins Before Birth (B4) Study at Massachusetts General Hospital (MGH) aims to discover, develop, and introduce novel interventions that protect the developing fetal brain through the implementation of a “learning” birth cohort. Our tripartite, multidisciplinary approach involves (1) deeply phenotyping the prenatal environment of risk and resilience for psychopathology through maternal surveys administered at multiple points during and immediately following pregnancy; (2) measuring early neurodevelopmental outcomes through the first years of life; and (3) triangulating exposure and outcome data with underlying biological mechanisms that can be harnessed to refine intervention strategies.

## Methods

### Study Aims

The B4 Study aims to canvass the prenatal environment for biological, psychosocial, and environmental stressors that associate with poorer early-childhood neurodevelopmental indices and may ultimately predict risk for neuropsychiatric illness. Further, the B4 Study aims to discover protective elements of the prenatal environment that mitigate risk conferred by environmental stressors on subsequent risk for neuropsychiatric sequelae.

### Stakeholder Engagement & Study Conception

The idea for the B4 Study as a learning birth cohort was conceived first through the engagement of key departmental stakeholders, starting with the president of MGH and followed by the chiefs of the departments of Psychiatry, Obstetrics and Gynecology (OB/GYN), Pediatrics, Neonatology, and Internal Medicine. The B4 Study was developed in tandem with Mass General Neuroscience, a flagship initial cross-departmental research project, and as such, the core B4 leadership team includes experts in neuroimaging and translational neuroscience research. As the B4 cohort matures, we plan to expand data collection to include neuroimaging and genomics; with its state-of-the-art facilities, MGH is well-positioned to accommodate these future directions.

### Study Setting & Population

Over 3600 births occur yearly at MGH’s main campus in downtown Boston^60^. MGH is also home to a sociodemographically diverse patient population enriched by multiple community-based clinics throughout the Boston area, with over 140 nationalities and 128 languages represented. Around 80% of all prenatal care visits take place at the MGH main campus in downtown Boston, with the remaining 20% at community health centers in Chelsea, Revere, and Charlestown. The community health centers – especially the one in Chelsea, where nearly 70% of residents are Hispanic or Latine and just under half are born outside of the United States – contribute significantly to the sociodemographic variation within the MGH patient population, offering a broader range of backgrounds and experiences compared to the main campus.^61^

### Study Design

The B4 Study is a prospective birth cohort study recruiting pregnant patients at MGH and continuing through the first 2 years of life. Created during the pandemic, the B4 Study can be completed entirely remotely and consists of self-reported surveys administered up to 3 times during pregnancy; once within 2 weeks postpartum; once approximately 1-2 months postpartum; and every six months of the child’s life for the first two years. A schematic of the study design is shown below (Figure 1).

**Figure 1.**
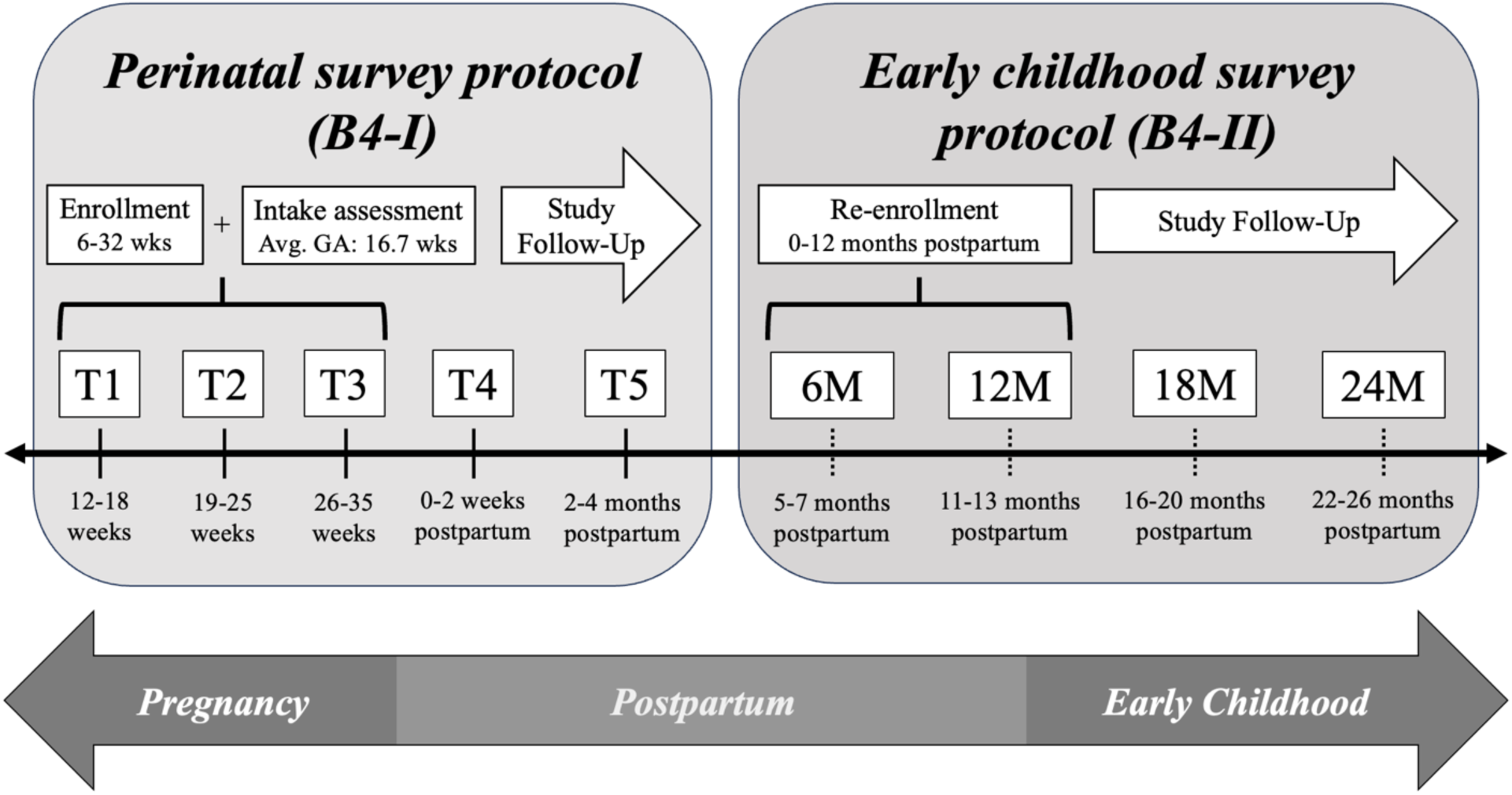
**B4 Study Assessment Timing.**

### Eligibility Criteria

Participants in the B4 Study must be 18 years or older; able to read or write in English or Spanish; pregnant at the time of recruitment; and receiving prenatal care at MGH. Patients were not enrolled in the study if they were above 34 weeks gestation, and the study team waited to make contact until they were at least 6 weeks gestation due to a significantly elevated risk for fetal loss before this point.^62^ Aside from the above, there are no additional exclusion criteria for the B4 Study.

If a participant becomes ineligible for the study during their pregnancy or after the birth of their child, they are removed from the study. The most common reasons for ineligibility include fetal or infant loss and transfer of prenatal care.

If participants complete at least one of the three scheduled timepoint surveys during their pregnancy, they are eligible to receive the two postpartum surveys about their childbirth experience and their infant’s early development. Participants are then contacted to join the pediatric follow-up study (B4-II), and verbal consent is obtained over the phone. Participants who do not complete any surveys during their pregnancy are not eligible to join the pediatric follow-up study.

### IRB Protocols & Consent Procedures

All study procedures and recruitment documents were approved by the Massachusetts General Hospital Institutional Review Board (IRB) prior to the start of the study. IRB approval was granted for electronic consent (eConsent) via Research Electronic Data Capture (REDCap) software hosted at Mass General Brigham. REDCap is a secure, web-based platform designed to support data capture for research studies.^63,64^ For participants who were not able to complete eConsent, verbal consent was also approved by the IRB, where staff explained details of the study over the phone and asked for verbal confirmation of the patient’s consent to participate. All participants were given the chance to ask questions about the B4 Study, address any concerns, and/or decline participation.

### Training of Study Staff

Upon joining the team, members of the study staff were required to complete standard hospital training in compliance with Health Insurance Portability and Accountability Act (HIPAA) guidelines; clinical research conduct; good clinical practice; human subjects research, and Epic electronic health records software. Staff members were expected to renew this training every three years. In addition, staff members received ongoing training in the IRB-approved protocol and consent procedures.

### Recruitment & Retention

In October 2020, details about the B4 Study were presented at an OB/GYN practice meeting, and five linked prenatal care providers (3 general OB/GYNs, 1 maternal-fetal medicine physician, and 1 certified nurse midwife) agreed to assist the study staff with recruitment in the MGH outpatient obstetrics clinic. Between January 2021 and December 2022, providers introduced the study to all eligible patients coming in for an initial prenatal care visit. Potential participants are identified via a Department Appointments Report on Epic, which lists all patients scheduled for an initial prenatal care visit with any of the five provider partners. Personalized recruitment materials are then delivered to each potential participant, and 1-2 weeks later, a member of the study staff attempts contact via phone (Figure 2).

**Figure 2.**
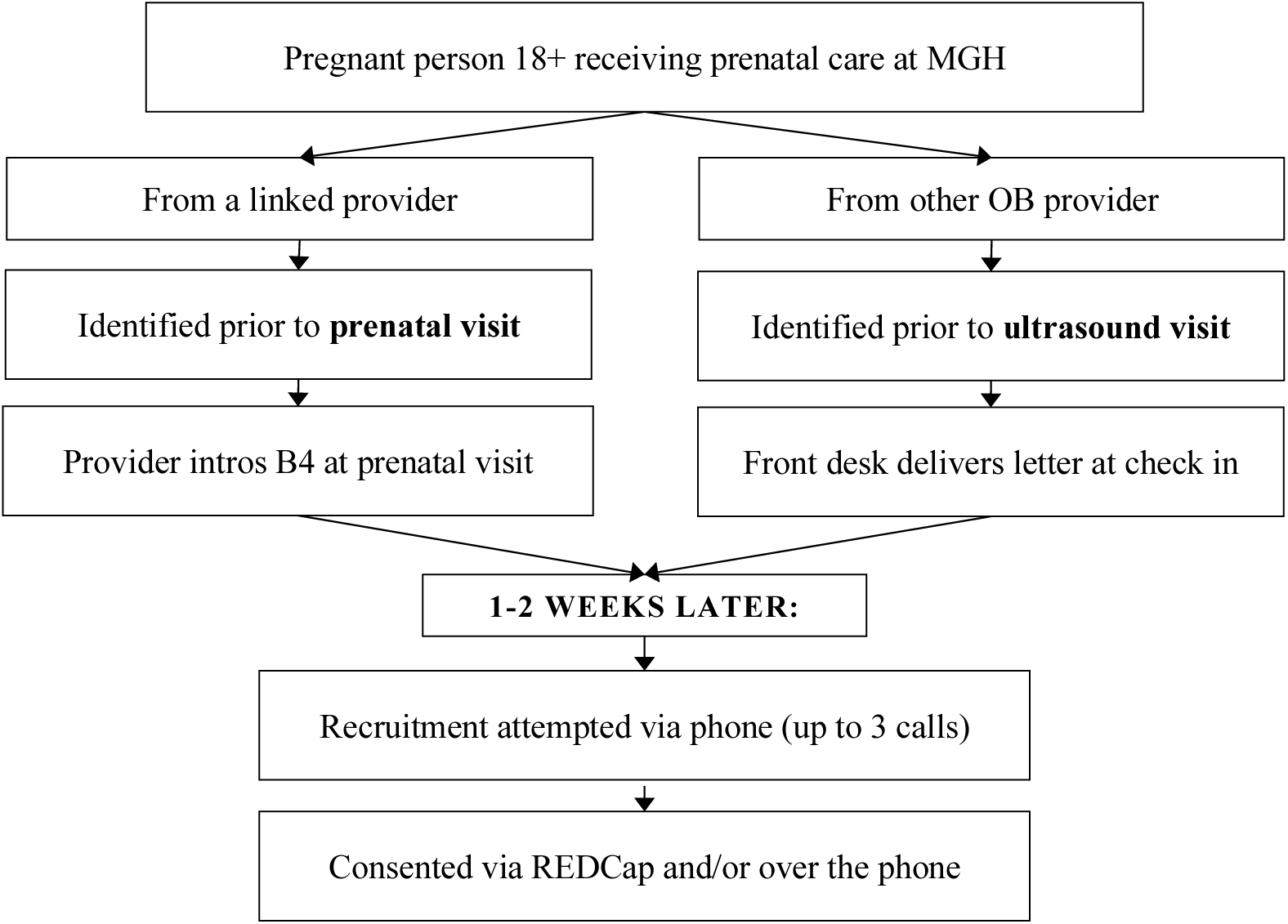
**B4 Study Recruitment Schematic.**

Beginning in July 2021, recruitment expanded to include patients coming in for ultrasound appointments, which serve not only those receiving prenatal care at MGH but also patients from community health centers in Chelsea, Revere, and Charlestown, providing access to a broader and more diverse patient population. In ultrasound recruitment, a list of eligible participants is given to the front desk staff at the MGH outpatient obstetrics clinic enclosed with recruitment letters for each patient listed (Figure 2). Additionally, participants can opt to be contacted by the study team via Rally, a database developed by Mass General Brigham Research to advertise research studies to potential participants.

Once eligibility criteria are confirmed by a member of the study staff, informed consent is obtained either electronically via REDCap or verbally over the phone. Participants are then entered into the B4 Study REDCap database and sent the intake survey, as well as the timepoint survey corresponding to their gestational age. To increase study retention, participants are sent three reminders in the two-week window following the receipt of each survey invitation.

### Measures and Data Collection

At the time of enrollment in the B4 Study, participants are asked to complete a brief intake survey assessing sociodemographics such as race, ethnicity, income level, and educational attainment; history of mental and physical health conditions; pregnancy intention; trauma history; and perceptions of neighborhood safety, social cohesion, and personal resilience (Appendix 1). Following the intake survey, each timepoint survey consists of up to 6 named sections, such as *Your Moods and Feelings*, *Your Daily Routines*, *Your Experiences With COVID-19*, and *Your Healthcare Experiences During Pregnancy*, among others (Appendix 2). Timepoint surveys in B4-I take between 15 and 90 minutes to complete.

Participants enrolled in the pediatric follow-up study (B4-II) are asked to complete timepoint surveys every 6 months for the first two years of life. B4-II surveys are also comprised of 6 sections, including *Your Background*, *Your Daily Routines*, *Your Mental Health*, *Your Relationships*, *Your Child’s Health*, and *Your Child’s Development* (Appendix 3).

### Medical Record Review

Thorough reviews of the birthing parent’s medical chart in Epic are conducted intermittently throughout their pregnancy to ensure continued fetal viability. After the participant gives birth to their child, a member of the study staff reviews their medical chart to assess prenatal care utilization; document any conditions or events complicating pregnancy and/or childbirth; and document relevant vital signs of the birthing parent and the newborn, including but not limited to birthweight, head circumference, duration of labor, and birth method. In addition, charts are reviewed for certain risk and resilience factors in the perinatal and periconceptional period such as pregnancy intention, folic acid supplementation, substance use, and social determinants of health (Appendix 4).

### Accessibility

The following steps were taken to ensure the inclusion of participants regardless of literacy, language, or steady internet access. All study materials and forms were written at a fifth-grade reading level and provided in English and Spanish. Throughout the study’s duration, at least 1-2 clinical research coordinators fluent in Spanish were employed at any given time. Participants were also given various options for survey completion, including online via REDCap; over the phone; or mailed and completed via pen and paper (or any combination of online, phone, and mail).

### Enrollment in Other Studies

Consistent with the initial goal of the B4 Study as a learning cohort, several observational add-on studies have been offered to the cohort, which will be described in subsequent papers. Studies included biosensor pilot studies and qualitative studies featuring both patients and providers.

## Discussion

Characterizing psychopathology risk is vital to understanding the complex landscape of early brain development; however, with so many risk factors being relatively unmodifiable—such as genetic and sociodemographic factors—presenting risk without resilience leaves families and providers without tangible next steps. Through the evolution of the B4 learning cohort, modifiable resilience factors will be identified and prospectively tested, linking perinatal behaviors and exposures to variation in maternal and child health outcomes. In addition, the learning cohort model allows for ongoing incorporation of participant feedback and consistent improvement of the study design to meet participants’ needs.

To maximize reproducibility and generalizability of study findings, a diverse and engaged participant population is crucial. Expanding recruitment efforts to ultrasound appointments and thereby including patients who receive care at affiliated community clinics has been one of many efforts to ensure a patient population representative of the diversity of the Boston area.

Additionally, bilingual research staff and accessible study materials in English and Spanish helped facilitate participation across varying literacy levels and language preferences. Recruitment, retention, and engagement efforts in community health centers in Chelsea, Revere, and Charlestown will remain a priority as the study progresses.

Additionally, the findings of the present study are limited to the validity of patient self-report, introducing potential reporting bias. While triangulation of self-reports with extensive medical record review will enhance study findings’ accuracy and reliability, the impact of reporting bias will be considered when interpreting the study results.

The B4 study represents a significant opportunity to enhance understanding of early risk and resilience factors to child psychopathology and address critical gaps in preventive psychiatry. To address limitations and improve study design, ongoing efforts to expand recruitment, streamline data collection, and integrate participant feedback are pivotal. The learning cohort model not only facilitates continuous refinement of study methods but also promises to yield valuable insights that can inform more effective, targeted interventions, working to improve the lives and developmental trajectories of young people.

## Data Availability

Data that has been and will be generated by the present study may be made available upon reasonable request to the authors.

## Acknowledgments

We thank the MGH Obstetrics and Gynecology clinic providers and staff who supported study recruitment, as well as all past, present, and future participants in the B4 Study.

## Appendix 1. Constructs Included in the B4 Study Intake Survey.

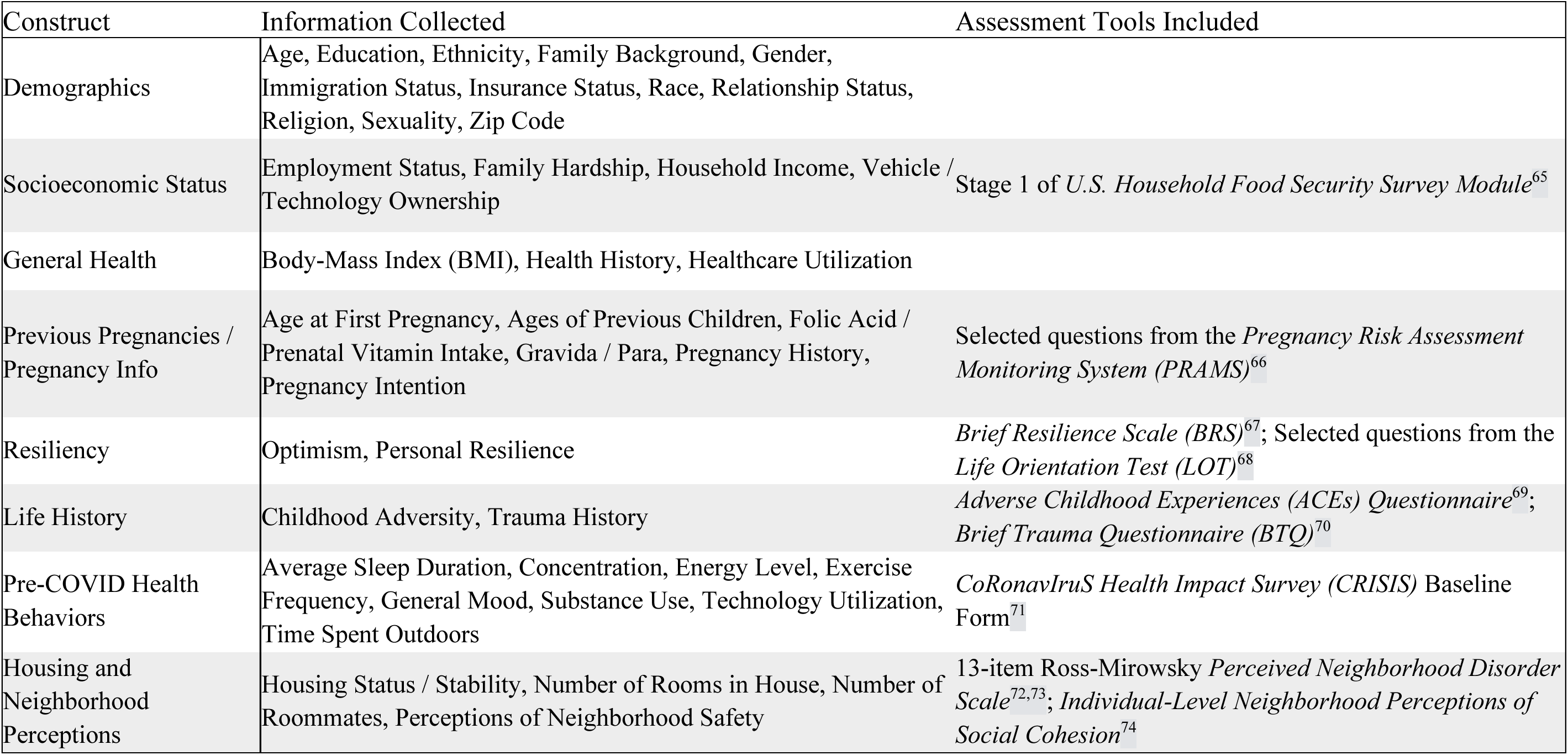

## Appendix 2. Sections Included in Each of the B4-I Study Timepoint Surveys.

| Section Title | Timepoints | Information Collected | Assessment Tools Included |
| --- | --- | --- | --- |
| Your Daily Routines | T1, T2, T3 | Alcohol and Substance Use | Consumption questions from the *Alcohol Use Disorders Identification Test (AUDIT-C)*<sup>75</sup>; *Drug Abuse Screening Test (DAST-10)*<sup>76</sup>; *Tobacco, Alcohol, Prescription medications, and other Substance use (TAPS)*<sup>77</sup> |
|  |  | Exercise / Physical Activity | Questions from the *Nurses' Health Study 2 (NHS-II) 2017 Long Form*<sup>78</sup> |
|  |  | Experiences of Discrimination | *Everyday Discrimination Scale (EDS)*<sup>79</sup> |
|  |  | Nutrition / Eating Habits | *Mediterranean Eating Patterns for Americans Screener (MEPA)*<sup>80</sup> |
|  |  | Sleep Behavior / Quality | *Pittsburgh Sleep Quality Index (PSQI)*<sup>81</sup> |
|  |  | Social Determinants of Health | Questions from the *Boston Medical Center THRIVE Screening Tool*<sup>82</sup> |
| Your Moods and Feelings | T1, T2, T3 | Anxiety | *Generalized Anxiety Disorder (GAD-7)*<sup>83</sup> |
|  |  | Gratitude | *Gratitude Questionnaire – 6-item form (GQ-6)*<sup>84</sup> |
|  |  | Mindfulness | *Cognitive and Affective Mindfulness Scale – Revised (CAMS-R)*<sup>85</sup> |
|  |  | Perinatal Depression | *Edinburgh Postnatal Depression Scale (EPDS)*<sup>86</sup> |
|  |  | Social Isolation | *Patient-Reported Outcomes Measurement Information System (PROMIS) Item Bank v2.0 – Social Isolation*<sup>87</sup> |
| Your Relationships | T1, T2, T3 | Intimate Partner Violence | *Ongoing Abuse Screen (OAS)*<sup>88</sup> |
|  |  | Relationships with Healthcare Team | *Discrimination in Medical Settings (DMS) Scale*<sup>89</sup>; *Provider Trust Scale (PTS)*<sup>90,91</sup> |
|  |  | Social Satisfaction | *Couple Satisfaction Index (CSI)*<sup>92</sup>; *Family Satisfaction Scale (FSS)*<sup>93</sup> |
|  |  | Social Stress | *Massachusetts Pregnancy Risk Assessment Monitoring System (PRAMS) Phase 8*; Selected Questions from the *COVID-19: Supporting Parents and Children during the Epidemic (Co-SPACE) Study Survey*<sup>94</sup> |
|  |  | Social Support | *Berkman-Syme Social Network Index*<sup>95</sup> |
| Your Experiences with COVID | T1, T2, T3 | Material / Financial Impact of COVID | *The CoRonavIruS health Impact Survey (CRISIS)*<sup>71</sup>; *Epidemic-Pandemic Impacts Inventory* |
|  |  | Mental Health Impact of COVID | *Abbreviated PTSD Checklist – Civilian Version*<sup>96</sup>; *Cohen's Perceived Stress Scale*<sup>97</sup>; *Depression Anxiety Stress Scale*<sup>98</sup>; *Impact of Event Scale – Revised 6-Item Survey*<sup>99</sup> |
|  |  | Social Impact of COVID-19 | *Medical Outcomes Study Social Support Survey*<sup>100</sup>; *The CoRonavIruS health Impact Survey (CRISIS)*<sup>71</sup> |
| Your Prenatal Care and Health | T3 | Prenatal Health and Risk | *Pregnancy Risk Assessment Monitoring System (PRAMS)*<sup>66</sup> |
| Your Upcoming Childbirth | T3 | Breastfeeding Planning | *Index of Breastfeeding Status*<sup>101</sup> |
|  |  | Expectations of Childbirth / Fear of Childbirth | *Wijma Delivery Expectations Questionnaire*<sup>102</sup> |
|  |  | Maternal Attachment | *Attachment Fraley Questionnaire*<sup>103</sup> |
|  |  | Personality | *Personality NEO Five Factor Inventory*<sup>104</sup> |
|  |  | Trauma History / Trauma Related to Pregnancy and Forthcoming Birth | *Brief Symptom Inventory (BSI)*<sup>105</sup>; *Life Event Checklist*<sup>106</sup>; *Childbirth PTSD Checklist*<sup>107–109</sup>; *Posttraumatic Growth Inventory (PTGI) - Pregnancy*<sup>110</sup>; *Civilian PTSD Checklist*<sup>107</sup> |
| Your Recent Childbirth | T4, T5 | Breastfeeding Status | *Index of Breastfeeding Status*<sup>101</sup> |
|  |  | Childbirth Experience / Perinatal Mental Health | *Edinburgh Postnatal Depression Scale (EPDS)*<sup>86</sup>; *Wijma Delivery Experiences Questionnaire*<sup>102</sup>; *Brief Symptom Inventory*<sup>105</sup>; *Peritraumatic Distress Inventory*<sup>111</sup>; *Peritraumatic Dissociative Experiences Questionnaire*<sup>112</sup>; *Childbirth PTSD Checklist*<sup>107–109</sup>; *PTGI - Childbirth*<sup>110,113</sup> |
|  |  | Infant Development | *Ages and Stages Questionnaire*<sup>114</sup> |
| Your Healthcare Experience During Pregnancy | T5 | Experiences with Healthcare Providers During Pregnancy, Labor, and Delivery | *Discrimination in Medical Settings Scale*<sup>89</sup>; *Provider Trust Scale*<sup>90,91</sup> |

## Appendix 3. Sections Included in Each of the B4-II Timepoint Surveys.

| Section Title | Timepoints | Information Collected | Assessment Tools Included |
| --- | --- | --- | --- |
| Your Background | 6M, 12M, 18M, 24M | Insurance Status, Relationship Status, Employment Status |  |
| Your Background | 6M, 12M, 18M, 24M | Housing, Neighborhood and Community Context | 13-item Ross-Mirowsky *Perceived Neighborhood Disorder Scale*<sup>72,73</sup>; *Individual-Level Neighborhood Perceptions of Social Cohesion*<sup>74</sup>; *Family Resources Scale – Revised*<sup>115</sup>; *Perceived Housing Stress*<sup>116</sup> |
| Your Daily Routines | 6M, 12M, 18M, 24M | Nutrition / Eating Habits | *Mediterranean Eating Pattern for Americans Screener (MEPA)*<sup>80</sup>, *Infant Feeding Practices*<sup>117</sup> |
|  |  | Food Insecurity | *Household Food Insecurity Access Scale Version 3*<sup>118</sup> |
|  |  | Alcohol and Substance Use | Consumption questions from the *Alcohol Use Disorders Identification Test (AUDIT-C)*<sup>75</sup>; *Drug Abuse Screening Test (DAST-10)*<sup>76</sup>; *Tobacco, Alcohol, Prescription medications, and other Substance use (TAPS)*<sup>77</sup> |
|  |  | Sleep Behavior / Quality | *Pittsburgh Sleep Quality Index (PSQI)*<sup>81</sup> |
| Your Feelings and Emotions | 6M, 12M, 18M, 24M | Perinatal Mental Health | *Edinburgh Postnatal Depression Scale (EPDS)*<sup>86</sup>, *Postpartum Distress Measure*<sup>119</sup> |
| Your Relationships | 6M, 12M, 18M, 24M | Social Satisfaction & Support | *Family Satisfaction Scale (FSS)*<sup>93</sup> *Couple Satisfaction Index (CSI)*<sup>92</sup>; *Medical Outcomes Study Social Support Survey (MOS-SS)*<sup>100</sup>; *Brief Attachment Scale (BAS-16)*<sup>120</sup> |
| Your Child's Health | 6M, 12M, 18M, 24M | Child's Behavior, Physical Health and Healthcare Accessibility | *National Survey of Children's Health A: This Child's Health*<sup>121</sup> |
| Your Child's Development | 6M, 12M, 18M, 24M | Child's Behavior | *Baby Pediatric Symptoms Checklist*<sup>122</sup>; *Ages and Stages Questionnaire*<sup>114</sup> |

## Appendix 4. Variables Included in B4 Participant Chart Review.

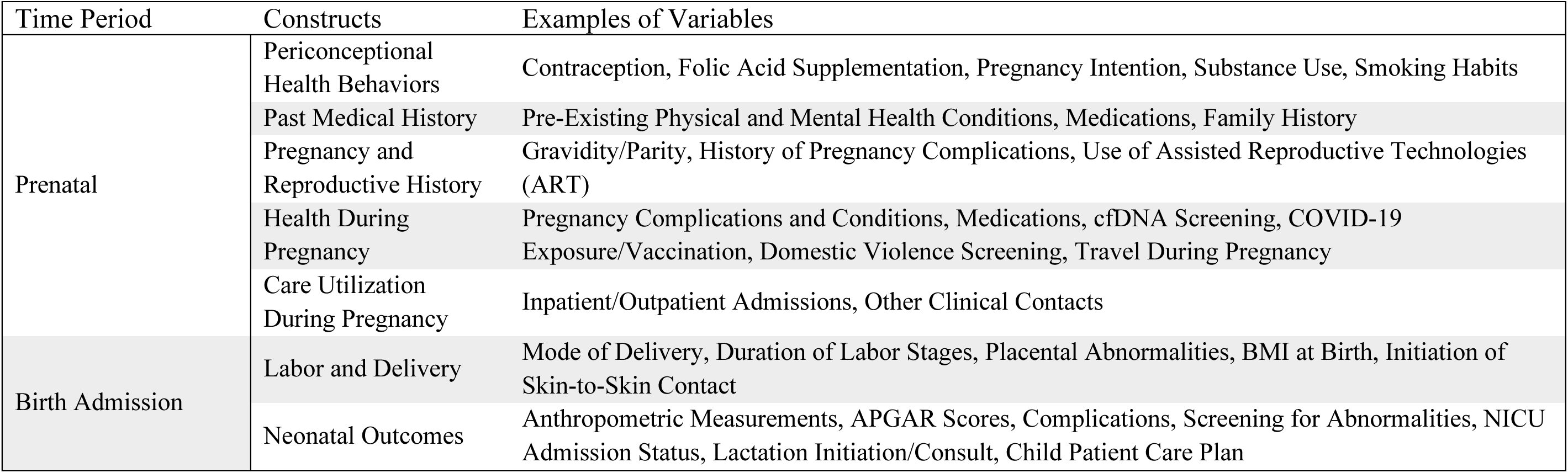

